# Molecular characterization of non-small cell lung cancer tumors in Latin American patients from Brazil, Chile and Peru uncovers novel potentially driver mutations

**DOI:** 10.1101/2020.09.11.20171025

**Authors:** Gonzalo Sepúlveda-Hermosilla, Alejandro Blanco, Matías Freire, Rodrigo Lizana, Javier Cáceres-Molima, Diego Ampuero, Paola Pérez, Liliana Ramos, Osvaldo Aren, Sara Chernilo, María Loreto Spencer, Jacqueline Flores, Giuliano Bernal, Mónica Ahumada Olea, Germán Rasse, Carolina Sánchez, Katherine Marcelain, Solange Rivas, Maria Galli de Amorim, Gabriela Branco, Diana Noronha Nunes, Emmanuel Dias-Neto, Helano C. Freitas, Cristina Fernández, NIRVANA Team, Rodrigo Assar, Ricardo Armisén

## Abstract

**Introduction:** Therapies that target activating *Egfr, Alk, Ros1* and other mutations have become first-line treatments that improve NSCLC patient’s life expectancy. Latin-American patients are poorly represented in clinical trials and in genomic databases, thus little is known about the prevalence of actionable mutations in this population. This study characterizes, for the first time, the somatic mutations found in 52 actionable genes, and describe a novel set of potentially actionable mutations, in NSCLC patients from Chile, Brazil and Peru, while correlating these genomic occurrences with relevant clinical, demographic and pathology aspects.

**Methods:** 1732 subjects diagnosed with NSCLC were analyzed. DNA and RNA were sequenced using a 52 genes NGS panel. Mutations were annotated using the Variant Effect Predictor, COSMIC, OncoKB and the Cancer Genome Interpreter to categorize somatic mutations.

**Results:** We found a total of 1713 mutations with 626 (36.5%) novel, potentially driver mutations. 66.1% of these novel mutations were predicted as Tier 1 driver mutations. Actionable mutations for *Ret and Alk* were more prevalent in Brazil than in Chile, whereas *Met* exon-14 skipping was significantly enriched in Chile. In Peru, *Egfr* is higher while *Kras* is lower. A high number of novels potentially driver mutations in know NSCLC actionable genes, such as *Alk, Erbb2, Ret, Met*, and *Ros1*, was found.

**Conclusions:** The analysis of many Latin America subjects revealed a significant number of clinically actionable but also novel somatic mutations in cancer genes highlighting the importance of including less-represented populations in clinical trials and molecular studies.

## Introduction

In 2018, lung cancer was the world’s most common cancer, both in terms of new cases (2.1 million cases, 11.6% of total) and deaths (1.8 million deaths, 18.4%) with high mortality rates and associated to countries with low human-development index ^1, 2, 3^. Approximately 85% of lung cancer is defined histologically as non-small cell lung cancer (NSCLC). NSCLC usually is diagnosed at a non-resectable locally advanced stage (Stage IIIB) or metastatic (Stage IV), when no curative treatment is available. Although immunotherapy appears as an alternative, new therapeutic agents with novel mechanisms of action are still desperately needed^4, 5^.

Advances in precision medicine, where patients are treated with therapies directed against specific molecular alterations driving their cancers, have transformed oncology ^6^. In Europe and North America, thanks to global efforts such as The Cancer Genome Atlas (TCGA) and the International Cancer Genome Consortium (ICGC), the landscape of actionable genomic alterations is well known, and a comprehensive set of oncogenic clinically actionable mutations for NSCLC tumors has been vetted, opening the possibility of prescribing effective targeted therapies. These include gene rearrangements and mutations in *Egfr, Alk, Ros1, Erbb2, Met, Map2k1, Braf, and Ret*, which collectively are found in about a 64% of lung adenocarcinomas patients. Approximately 10 – 40 % of NSCLC patients show tumor-associated mutations in the Epidermal Growth Factor Receptor (Egfr)^7^. The identification of patients whose tumors have *Egfr* mutations, predicts the efficacy of EGFR inhibitors (i.e., exon 19 deletions, L858R and other *Egfr* activating mutations) ^8^. *Alk* gene fusions are reported in 3%-5% of patients. Basic and clinical research suggests that the frequency of the gene rearrangements that can be targeted by specific tyrosine kinase inhibitors is similar for Asian and Western patients ^9^. Chromosomal rearrangements involving *Ros1* were originally described in glioblastomas. For NSCLC, *Ros1* mutations, presents a frequency of around 6%, according to TCGA data. The fusion *Slc34a2-Ros1* was identified as a potential driver mutation in a NSCLC cell line (HCC78); meanwhile, *Cd74-Ros1* was found in a NSCLC patient sample ^10^. These fusions confer constitutive kinase activity to *Ros1* and are associated with *in vitro* and *in vivo* sensitivity to tyrosine kinase inhibitors.

Despite the relevance and recent availability of novel TKI drugs, little is known in Latin-American countries about the molecular epidemiology of clinically actionable mutations in the region. Available regional efforts are focused in few genes (such as *Egfr, Alk* and *Kras)* and testing methodologies used are not necessarily equal across different countries and cohorts ^11-15^. In addition, large cancer genomics efforts, like TCGA and ICGC, are severely deprived of minorities (including subjects of Hispanic ethnicity), limiting the capacity to describe somatic mutations with a prevalence below 10%, and to overcome the background of the somatic mutation frequency in specific ethnic groups^16^.

Here we present the results of a large harmonized NGS-based screening of the NSCLC actionable mutations in Latin-American patients (particularly enriched in Hispanic and admixed populations). We characterized novel and known molecular features of actionable genes for NSCLC in Brazil, Chile and Peru. Finally, to characterize actionable patients’ subgroups, we correlate the findings with relevant demographic, clinical and tumor aspects. This work aims to reduce the gap in cancer genomics diversity and disparities observed in large international cohorts and the current lack of ethnic diversity in global cancer databases^16, 17^, therefore supporting efforts to improve molecular diagnosis and treatment in Latin America, a region that is home to more than 648 million people^18^.

## Material and methods

### Study Design

The NIRVANA protocol (Validation of Molecular Diagnostic Technologies for Lung Cancer Patients) was approved by local ethics committees for each recruiting hospital in Chile, Brazil and Peru, and registered at clinicaltrials.gov as NCT03220230. From July 2015 to October 2018, patients 18 years old or older, with a histological or cytological diagnosis of NSCLC consented to contribute FFPE lung cancer tissue samples, along with demographic, clinical, and pathology information. Primary or metastatic NSCLC samples were collected as part of the standard of care biopsy procedures as indicated by treating physicians. Up to 8 FFPE 5 μm sections with at least 5% of tumor cellularity were selected for isolation of RNA and DNA, using the *RecoverAll* extraction kit (Thermo Fisher Scientific).

### Sequencing quality control criteria

Libraries were prepared using Oncomine Focus Assay (OFA, Thermo Fisher Scientific) and sequenced in the Ion Personal Genome Machine System. OFA is a targeted NGS panel, to discover SNVs, Indels and CNVs in the DNA of 35 actionable genes, and actionable gene fusions in 23 genes in the tumor RNA. The quality control metrics thresholds were set as at least 200.000 reads and 80% of On-Target reads for DNA libraries, and 5.000 reads correctly mapped plus three out of five expression-control amplicons detected, for RNA.

### Alignment and Variant Calling

Analyses from mapping to variant calls were made using the Ion Reporter software v5.2 (Thermo Fisher Scientific). Reads were aligned to the human reference genome hg19, using the Torrent Mapping Alignment software (Torrent Suite v5.2). Variant calling was performed with the Torrent Variant Caller 5.2 using the workflow “Oncomine Focus for DNA Single Sample” included in the Ion Reporter Software, with stringent parameters for calling SNVs and Indels: minimum allele frequency of 4% (SNVs) and 7% (Indel), minimum coverage supporting a variant was 15x (SNVs and Indels), minimum coverage of the variant location is 50x, and the variant scores, represented as a Phred-scaled value of the reads supporting a variant above the minimum frequencies was 6 for SNVs and 20 for Indels. For fusions, reads were aligned to a synthetic reference spanning the break-point sequences targeted by the OFA panel. The workflow “Oncomine Focus for Fusion in single sample” was used with the following parameters: a 70% overlapping read alignment with the reference and 66% of exact matches and a minimum valid mapped read of 20X and 15X for fusions and expression controls, respectively.

### VCF Processing and MAF generation

DNA VCF’s were processed with VCFTools v1.1: multi-allelic sites were split and left-normalized using the hg19 human reference. All the remaining reference/reference sites, variants with Allele Frequency < 5%, and observed alternate allele < 10 reads were removed from the DNA VCF’s. For the RNA VCF’s, only fusions with more than 20 reads were kept. Then, all the VCF’s were transformed to MAF’s using vcf2maf ^19^ and variants were annotated with Variant Effect Predictor (VEP) v93 ^20^, OncoKB ^21^ and COSMIC v81^22^ databases. For data queries, analysis and visualizations, MAF files and its clinical information were loaded into a local implementation of cBioPortal (The Hyve) ^23^.

### Variants Classification

Germline variants were determined from VEP annotation and excluded from further analysis. Somatic mutations were grouped based on the NSCLC OncoKB’s levels of evidence^21^ and complemented with COSMIC annotations into 4 categories: mutations with level 1 to 3b were considered *clinically actionable* in NSCLC, mutations associated to drug resistance, categorized as R1 or R2 were classified as *resistance; Likely-Actionable* was defined for any actionable alteration reported in other cancer types. Level 4 or *Known* alterations were defined for those with no clinical evidence and with at least one COSMIC entry. Any other alteration (excluding passenger mutations) not reported in OncoKB and not found in COSMIC were classified as *Novel*. To predict cancer drivers mutations and deleteriousness among these *Novel* mutations, they were further assessed with Cancer Genome Interpreter (CGI) ^24, 25^ using the clinical significance predictions according to ASCO’s guidelines for interpretation^26^: Tier 1 characterizes variants of strong clinical significance, with FDA-approved therapy in NSCLC, Tier 2 describes variants with potential clinical significance: FDA-approved therapies but for others tumor types. Tier 3 were variants of unknown clinical significance, and Tier 4 variants were benign or likely benign. CGI’s annotation also provides the passenger mutation categorization ^27^. A reference subset of 31 novel variants was characterized in the radarplots with a gray shade superimposed. These variants were concordantly annotated as deleterious, highly damaging, and located over sensitive domains by all the annotation algorithms. Highly mutated samples were defined as those with 5 or more genes showing at least one DNA variant or RNA fusion. DNA mutations are represented as lolliplots excluding splice variants. The protein associated with the longer transcript for each gene was used for domain’s protein representation. Lolliplots and oncoprints were made using cBioPortal^23^ and the maftools^28^ package (R core v.3.5), respectively. Missense mutations of the top five most mutated genes were represented with a Circos plot generated by Table Viewer ^29^.

### Statistical Analyses

Univariate and multi-parametric logistic regressions of demographic and clinical factors over gene mutations were performed considering only clinically actionable variants, known or likely-actionable mutations, using a Fisher exact test with Odds Ratios estimation (95% C.I.) on Python v.3.6.4 using pandas and matplotlib libraries. All further analyses are considered with a union set of 1732 subjects, with either DNA *or* RNA QC-approved (Table 1), except for prevalence calculations were only DNA and RNA QC-pass samples and clinically actionable variants, known or likely-actionable mutations and Tier1 and Tier2 Novel mutations were considered. Confidence intervals were calculated by Agresti-Coull’s method^30^. Z-test was used to assess significant differences between gene’s prevalence, mutation enrichment between countries and variants class subgroups using the statsmodels package. cBioPortal was used to access NSCLC Memorial Sloan Kettering Cancer Center (MSKCC) study^31^ as a reference population, spanning non-synonymous mutations from *Egfr, Kras, Met, Pik3ca* and *Alk* in 860 subjects (http://bit.ly/38OtHM0).

**Table 1.** Summary of sample and demographic-clinical features per country.

|  | Brazil (%) | Chile (%) | Peru (%) | Total (%) |
| --- | --- | --- | --- | --- |
| **Number of Subjects** | 479 (27.7) | 1127 (65.1) | 126 (7.3) | 1732 |
| **Subject Age (years)** |  |  |  |  |
| 18-26 | 1 (0.2) | 2 (0.2) | 0 (0) | 3 (0.2) |
| 27-35 | 4 (0.8) | 9 (0.8) | 6 (4.8) | 19 (1.1) |
| 36-44 | 18 (3.8) | 19 (1.7) | 10 (7.9) | 47 (2.7) |
| 45-53 | 50 (10.4) | 100 (8.9) | 13 (10.3) | 163 (9.4) |
| 54-62 | 134 (28) | 240 (21.3) | 23 (18.3) | 397 (22.9) |
| 63-71 | 154 (32.2) | 360 (31.9) | 29 (23) | 543 (31.4) |
| 72-80 | 91 (19) | 303 (26.9) | 29 (23) | 423 (24.4) |
| 81-89 | 23 (4.8) | 85 (7.5) | 16 (12.7) | 124 (7.2) |
| 90 and more | 4 (0.8) | 9 (0.8) | 0 (0) | 13 (0.8) |
| Mean | 64.1 | 66.7 | 63.9 | 65.8 |
| SD | 11.0 | 10.7 | 14.6 | 11.2 |
| Range | 23-93 | 25-94 | 27-89 | 23-94 |
| **Gender** |  |  |  |  |
| Female | 243 (50.7) | 525 (46.6) | 66 (52.4) | 834 (48.2) |
| Male | 236 (49.3) | 602 (53.4) | 60 (47.6) | 898 (51.8) |
| **Ethnicity** |  |  |  |  |
| Asian | 8 (1.7) | 3 (0.3) | 0 (0) | 11 (0.6) |
| Black | 27 (5.6) | 0 (0) | 0 (0) | 27 (1.6) |
| Caucasian | 230 (48) | 21 (1.9) | 0 (0) | 251 (14.5) |
| Hispanic | 76 (15.9) | 1072 (95.1) | 24 (19) | 1172 (67.7) |
| Miscegenation | 65 (13.6) | 9 (0.8) | 102 (81) | 176 (10.2) |
| Native Indian | 2 (0.4) | 4 (0.4) | 0 (0) | 6 (0.3) |
| Other | 71 (14.8) | 18 (1.6) | 0 (0) | 89 (5.1) |
| **Histology** |  |  |  |  |
| LUAD | 408 (85.2) | 926 (82.2) | 112 (88.9) | 1446 (83.5) |
| LUSC | 41 (8.6) | 178 (15.8) | 6 (4.8) | 225 (13) |
| Others NSCLC | 30 (6.2) | 23 (2) | 8 (6.3) | 61 (3.5) |
| **Tobacco use** |  |  |  |  |
| Never | 96 (20) | 348 (30.9) | 72 (57.1) | 516 (29.8) |
| Current | 33 (6.9) | 91 (8.1) | 0 (0) | 124 (7.2) |
| Former | 218 (45.5) | 659 (58.5) | 31 (24.6) | 908 (52.4) |
| Unknown | 132 (27.6) | 29 (2.6) | 23 (18.3) | 184 (10.6) |
| **Family history of cancer** |  |  |  |  |
| Yes | 216 (45.1) | 240 (21.3) | 19 (15.1) | 475 (27.4) |
| No | 109 (22.8) | 231 (20.5) | 32 (25.4) | 372 (21.5) |
| Unknown | 154 (32.1) | 656 (58.2) | 75 (59.5) | 885 (51.1) |
| **QC Pass Samples** |  |  |  |  |
| DNA | 318 (66.4) | 714 (63.4) | 95 (75.4) | 1127 (65.1) |
| RNA | 406 (84.8) | 992 (88) | 97 (77) | 1495 (86.3) |

## Results

### Cohort description and clinical and genomic data

As part of the NIRVANA study (NCT03220230) a total of 1732 NSCLC subjects were analyzed in Chile, Brazil and Peru. Tissue samples were obtained at the time of diagnosis (before any systemic or radiation treatment) and were subjected to an NGS targeted sequencing assay for the detection of hotspots, SNVs and indels in 35 genes, and gene fusions in 23 genes for a combined total of 52 cancer actionable genes. Most samples were obtained in Chile (65.1%), followed by Brazil (27.7%) and Peru (7.3%), from a total of 21 cities and 37 clinical sites (Table 1 and Figure 1A). Patient age at the time of diagnosis (65.8 years) was similar for the three countries; and female and males were equally represented. Regarding ethnicity, the cohort is mostly composed of Hispanic subjects (67.7%) follow by Caucasian (14.5%) and miscegenation (10.2%). Ethnicities are not balanced across countries: in Chile predominates Hispanics (95.1%), in Brazil Caucasians (48%) and in Peru miscegenated (81%), as depicted in Table 1. Is important to notice that ethnicity is self-assigned. In the three countries, adenocarcinoma was the most common histology (83.5%) followed by squamous cell carcinoma (13%). Current and former smokers account for more than 60% of the cohort. In 49% of subjects, information about family history of cancer was available, and 55% of these reported positive familial history (Table 1). Regarding subjects’ pathological stage, most of them were diagnosed at stage IV in Chile, Brazil and Peru, however nearly half of the Peruvian cases did not report stage information on the clinical records (Figure 1B). After sequencing, a total of 1127 (65% out of 1732 subjects) DNAs and 1495 (86% of 1732 subjects) RNA samples met the NGS quality control thresholds (Table 1).

**Figure 1.**
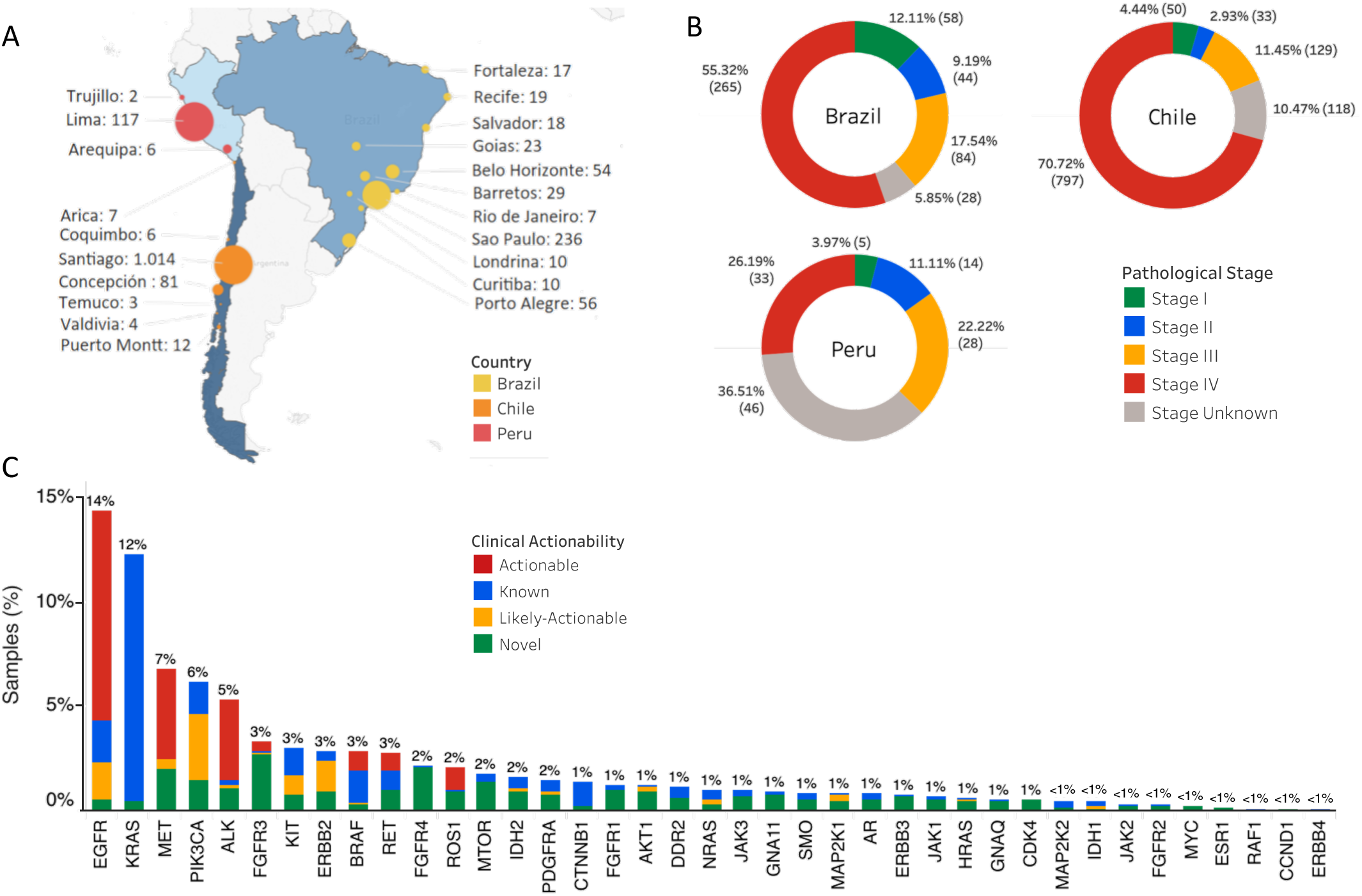
Cohort description - **(A)** Clinical sites along the three participating countries. The circle size represents the total subjects recruited in each city. Several clinical sites were aggregated at the city level. **(B)** Frequency of pathological stages per country. The pie charts show the proportion of the pathological stages of the samples at diagnosis in each country. **(C)** Percentage of samples altered per gene. Colors shows the clinical actionability of the variants detected in each gene.

Overall, 746 out of 1731 subjects, displayed at least one variant (SNVs, indel or fusions) in any of the 52 analyzed genes. The genes with the highest prevalence of mutations (actionable, likely actionable, without clinical actionability (known) and novel tier 1-2) in the NIRVANA cohort are *Egfr, Kras, Met, Pik3ca* and *Alk*, which account for 44% of all variants detected (Figure 1C). To comprehensively describe tumor somatic variants (SNVs, indels and gene fusions) and clinical and pathological information at subject level, waterfall plots were built for each cohort of subjects in each country (Supplementary Figure 1A-C for Chile, Brazil and Peru, respectively). The analysis focused on the most frequently mutated genes and NSCLC actionable genes. Most patients show a predominant mutated driver gene, however there are 41 patients whose tumors have mutations in multiple genes and a high mutational load: 12 (2.5%) are from Brazil, 27 (2.4%) from Chile, and 2 (1.6%) from Peru (Supplementary Figure 1A-C). Overall, these samples are significantly enriched in former smoker patients. The mutual exclusiveness analysis (Supplementary Figure 2) shows that *Egfr* mutations correlate negatively with *Alk, Erbb2, Fgfr3, Map2k1, Met, Pik3ca, Ret*, and *Ros1. Alk* also correlates negatively with *Erbb2, Met* and *Pik3ca*. Most significant co-occurrences are pairs involving low frequency genes like *Hras* with *Nras, Idh2* with *Kit, Idh2* with *Pdgfra*, and *Kit* with *Pdgfra*.

### Oncogene mutation’s prevalence per country

Although in Brazil, Chile and Peru Egfr, Kras, Pik3ca, Met and Alk accounts for around 50% of the mutations, proportions of mutated genes were distinctive for each of the analyzed countries. (Figure 2A). It is also interesting to note that the proportion of non-mutated subjects is different across countries: 29.9% on Brazil, 39.7% in Chile and 36% in Peru. Regarding low prevalence genes (<4%), the presence of NSCLC actionable ones such as Braf, Ret and Erbb2 reveal therapeutic opportunities in the three analyzed countries (Figure 2A).

**Figure 2.**
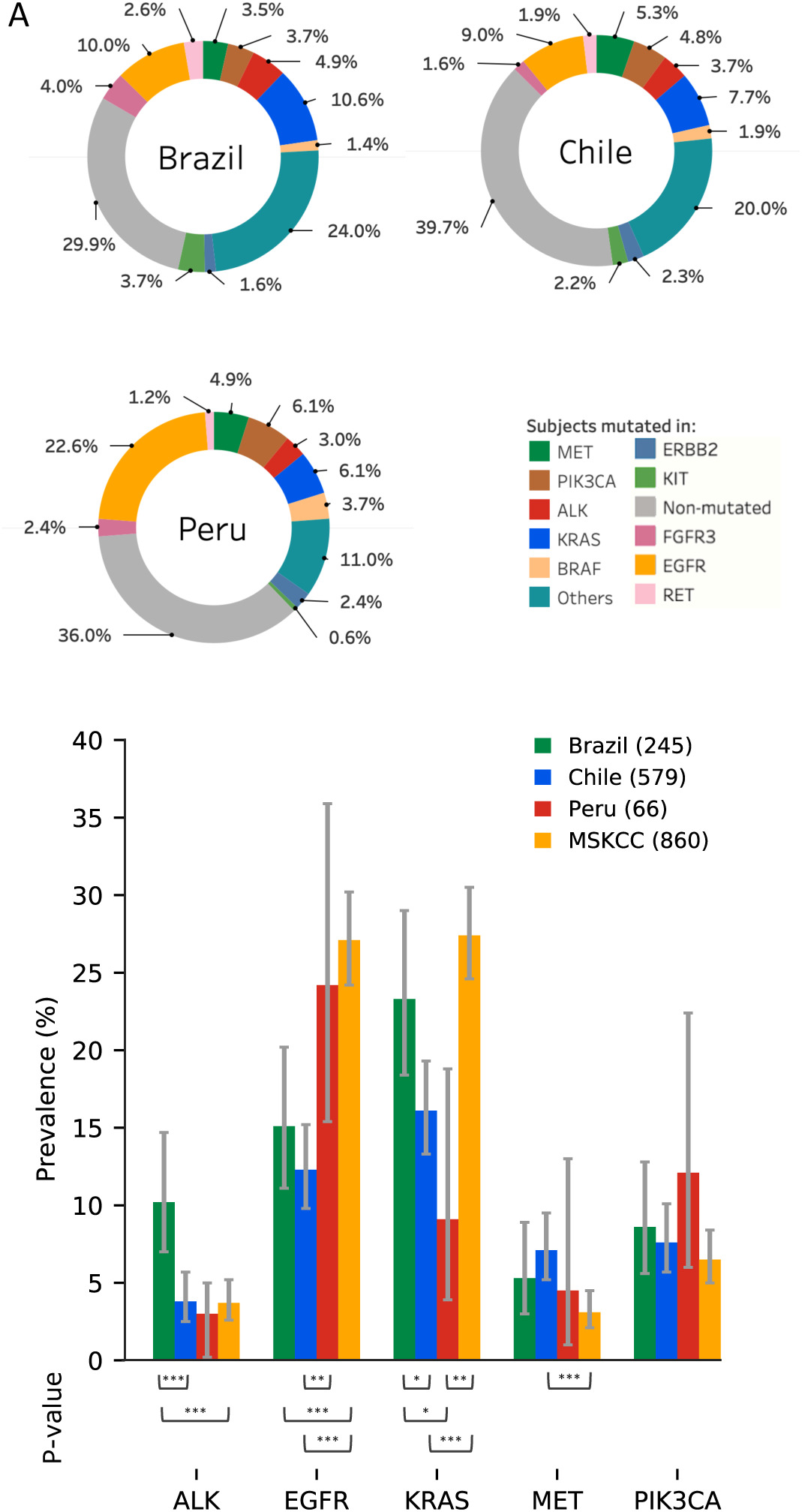
Description of alterations per country. **(A)** Pie charts describe the proportion of subjects in each country spanning overall alterations in the sequenced genes. Genes altered in less than 3% of the subjects in each country were aggregated as *Others*. **(B)** Colored bars show the prevalence of the five most altered biomarkers in the cohort. Only samples that passed QC in both DNA and RNA were considered, while passenger and non-protein affecting alterations were excluded from the analysis. 95% confidence interval bars are shown as grey error bars. Pairwise z-test was used to evaluate significant differences between countries, the p-values were ranked as: *** ≤ 0.001; ** ≤ 0.01; * < 0.05.

Gene specific prevalence (excluding non-protein affecting and passenger mutations) in 891 subjects with DNA and RNA data (Figure 2B), shows that Brazilian *Alk* mutation’s prevalence is significantly higher than in Chile (10.2% vs. 3.8%), and in the MSKCC cohort (3.2%). When the analysis is limited only to *Alk* gene fusions, a 3.7% (CI95%: 1.5 – 3.4) prevalence is observed in Chile, 3.1% (CI95%: 0 – 7.7) in Peru, and 6.7% (CI95%: 4.2 – 9) in Brazil. Prevalence of mutations in *Egfr* was halved in Chile (12%) and Brazil (15%), compared to MSKCC (27%) and Peru (24%). On the other hand, *Kras* mutations prevalence is statistically similar between Brazil (23%) and MSKCC (27%), while significantly lower in Chile (16%) and Peru (9%); while prevalence of mutations in Met is significantly higher in Chile than in MSKCC. No significant differences were found for *Pik3ca*.

### Actionable somatic mutations

A total of 560 patients had one or more actionable or likely actionable mutations (SNVs, indels or gene fusion). Regarding gene fusions, a total of 215 actionable fusions were detected (Table 2 and Supplementary Figure 3). *Met* exon-14 skipping (MET-MET.M13M15) is the most frequent rearrangement, comprising 75 events (35% out of the 215 fusions), mostly from Chilean samples (OR=2.09, Table 2 and Supplementary file 1). Subjects harboring MET fusions are generally adenocarcinomas (86.4%) followed by squamous cells (11.9%). Most of them are at stage IV (57.6%), former (61%) and current (8.5%) smokers, female (56%) and male (44%) with a 65-year-old median age (Supplementary file 2). *Eml4-Alk* fusions were the second most prevalent, with a total of 72 event (33%), including 32 V1 (E13A20, 44.4%), 11 V2 (E20A20, 15.2%), and 20 V3a/b (E6a/bA20, 27.7%) variants. E13A20 *Eml4-Alk* fusions are particularly enriched in Brazil (OR=3.9, Table 2 and Supplementary file 1). Also, there is an enriched prevalence of *Alk* rearrangements towards female, younger, native Indian patients (Supplementary table 1). Regarding Ret fusions, 10 *Ki5b-Ret* fusions were found and are enriched in Brazil (OR=3.1, Table 2 and Supplementary file 1). Other less frequent fusions in *Ros1* are also detected, including *EZR-Ros1* and *CD74-Ros1*. Subjects of Caucasian or Hispanic ethnicities are more likely to harbor a Ros1 actionable mutation (Supplementary table 1).

**Table 2.** Actionable mutations per country. Variants that occur less than 5 times were aggregated as *Others*.

|  | n | Brazil n(%) | Chile n(%) | Peru n(%) |
| --- | --- | --- | --- | --- |
| **Met (83)** |  |  |  |  |
| MET-MET.M13M15 | 75 | 11 (2.7) | 60 (6.0) | 4 (4.1) |
| X1010_splice | 7 | 0 (0) | 4 (0.6) | 3 (3.2) |
| Others | 1 | 0 (0) | 1 (0.2) | 0 (0) |
| **Alk (77)** |  |  |  |  |
| EML4-ALK.E13A20 | 32 | 19 (4.7) | 10 (1) | 3 (3.1) |
| EML4-ALK.E6aA20 | 15 | 5 (1.2) | 10 (1) | 0 (0) |
| EML4-ALK.E20A20 | 11 | 1 (0.2) | 10 (1) | 0 (0) |
| EML4-ALK.E6bA20 | 6 | 2 (0.5) | 4 (0.4) | 0 (0) |
| EML4-ALK.E18A20 | 6 | 0 (0) | 6 (0.6) | 0 (0) |
| Others | 7 | 2 (0.4) | 4 (0.4) | 1 (1) |
| **Ret (15)** |  |  |  |  |
| KIF5B-RET (K15R12, K23R12) | 11 | 8 (1.9) | 3 (0.3) | 0 (0) |
| Others | 4 | 0 (0) | 4 (0.4) | 0 (0) |
| **Ros1 (25)** |  |  |  |  |
| EZR-ROS1 (E10R34, E10R35) | 9 | 5 (1.2) | 4 (0.4) | 0 (0) |
| CD74-ROS1 (C6R32, C6R34, C6R35) | 6 | 4 (0.9) | 2 (0.2) | 0 (0) |
| Others | 10 | 2 (0.4) | 8 (0.8) | 0 (0) |
| **Egfr (210)** |  |  |  |  |
| L858R | 73 | 22 (6.9) | 36 (5.0) | 15 (16) |
| E746_A750del | 47 | 14 (4.4) | 26 (3.6) | 7 (7.4) |
| EGFR-EGFR.E1E8.DelPositive.1 | 16 | 3 (0.7) | 12 (1.2) | 1 (1.0) |
| L747_A750delinsP | 8 | 0 (0) | 6 (0.8) | 2 (2.1) |
| L747_P753delinsS | 7 | 2 (0.6) | 3 (0.4) | 2 (2.1) |
| G719A | 6 | 2 (0.6) | 3 (0.4) | 1 (1.1) |
| E746_S752delinsV | 6 | 3 (0.9) | 3 (0.4) | 0 (0) |
| S768_D770dup | 5 | 2 (0.6) | 2 (0.3) | 1 (1.1) |
| Others | 37 | 10 (3.0) | 22 (3.1) | 5 (5.0) |
| **Braf (17)** |  |  |  |  |
| V600E | 16 | 4 (1.3) | 8 (1.1) | 4 (4.2) |
| Others | 1 | 0 (0) | 0 (0) | 1 (1.1) |
| **ErbB2 (24)** |  |  |  |  |
| Y772_A775dup | 12 | 1 (0.3) | 9 (1.3) | 2 (2.1) |
| Others | 13 | 4 (1.3) | 8 (1.2) | 1 (1.1) |

Regarding the most frequently mutated genes in the cohort, most actionable and likely actionable SNVs mutations are observed in *Egfr* and Pik3ca (Figure 3). *Kras* and *Egfr* contribute most of the known variant class (Figure 3). Regarding SNVs and indels, most of the actionable and likely-actionable variants are detected in *Egfr* (Table 2). The most common *Egfr* actionable mutation is exon 21 L858R, representing 34.8% of the *Egfr* mutations, particularly enriched in Peru (Table 2). Exon 19 *Egfr* E746_A750del accounts for 22.4%. Also, there are 16 (7.6%) occurrences of *Egfr-* Egfr.E1E8.Del, commonly known as *Egfr* vIII deletion, lacking exons 2-7 (Table 2). Twenty-one exon 19 indels mutations are present: L747_A750delinsP, L747_P753delinsS and E746_S752delinsV. Besides, *Egfr* resistance mutations are found: four subjects with the mutation T790M are present, one with D770_N771insGL and five with S768_D770dup for a combined occurrence of 4.8% of *Egfr* actionable mutations. Hispanics, Caucasians or never smokers with a family history of cancer are more likely to harbor an *Egfr* actionable mutation (Supplementary Table 1). Concerning other NSCLC actionable genes, BRAF V600E and ERBB2 Y772_A775dup were found in 16 and 12 subjects respectively. Other less frequent mutations (<5 occurrences) are described in Supplementary file 1.

**Figure 3.**
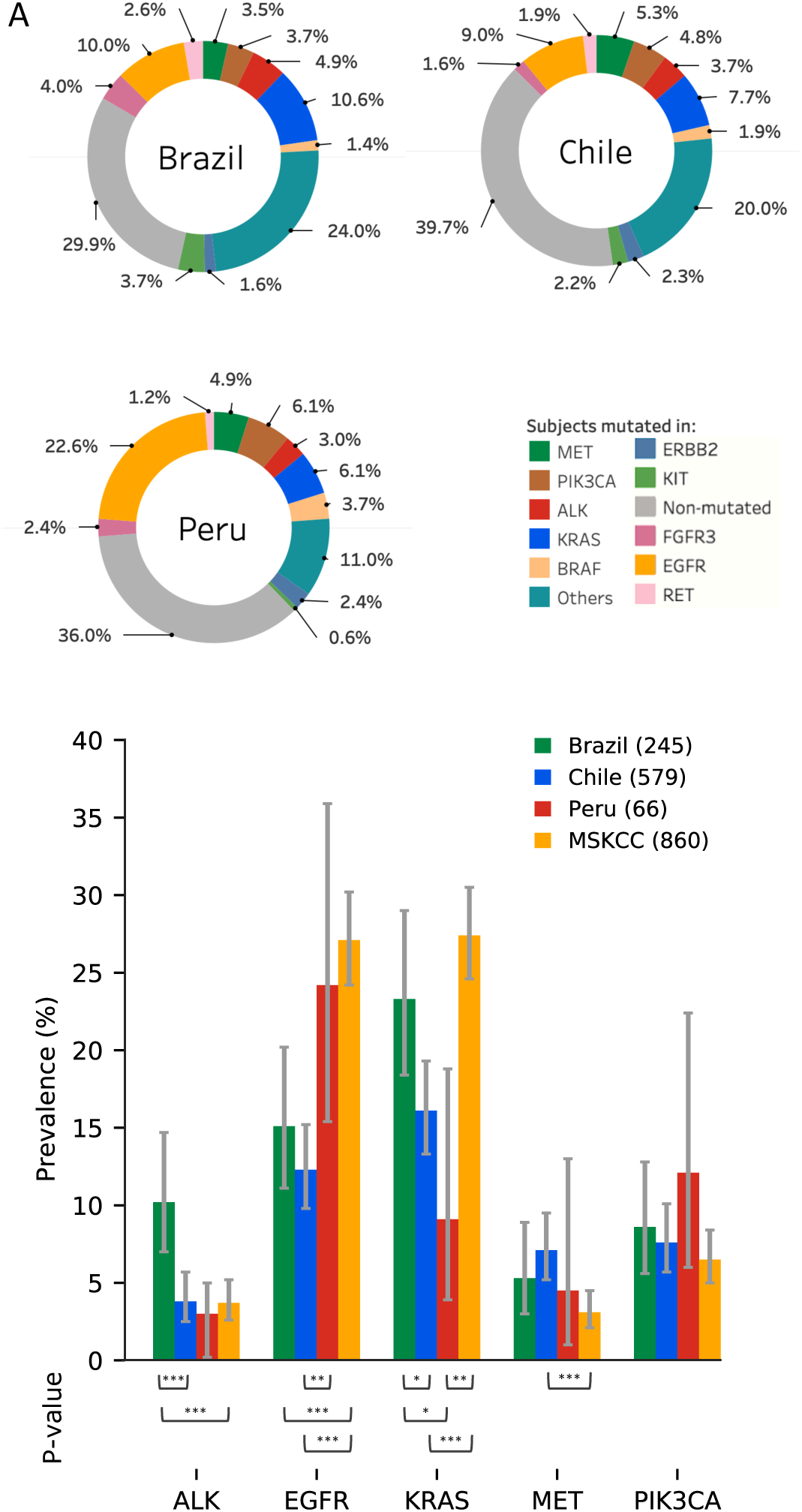
Landscape of missense variants and mutation class features. **(A)** Circos plot of the top 5 most mutated genes along with their clinical actionability. The most external ring color codes each variant which is linked to a specific type of mutation: actionable as red, likely-actionable as orange, known as blue, and novel as green. The width of the edges represents the proportion of the pair observed in the cohort. (**B)** 5-axis profile of relevant clinical aspects for each type of variant. *Females* axis represents the percentage of females samples, *Age* characterize the mean value of subjects ages, *Surgical Biopsies* aggregates the fraction of invasive biopsies (i.e. tumor resections and mediastinoscopy) against non-invasive biopsies (i.e. fibrobronchoscopies and percutaneous punctures), The *Non-Smokers* axis informs the fraction of non- or former smoker subjects. The *FHOC* axis shows the fraction of subjects having any reported relative with a history of cancer. The sixth axis displays the average CADD score, as a measure of the pathogenicity of the variants. The gray shade superimposed in each radar plot depicts the same 5-axis profile, selecting the 31 most relevant predicted driver novel variants. Asterisk on axis labels denotes significant differences (p-value < 0.05)

### Novel Variants

From a total of 1713 mutations, 626 (36.5%) of them were labeled as Novels (see materials and methods). The country with the highest number of novel mutations is Brazil with 232 (39.9% of its mutations), followed by Chile with 376 (36.6%), and Peru with 18 (17.1%). The genes with the highest proportion of novel mutations (more than 90%) are *Cdk4, Myc, Ccnd1, Esr1*, and *Fgfr4*. Among the group of NSCLC actionable genes, *Met* (90%), *Map2k1* (62.5%), *Ros1* (61.1%), *Alk* (59.5%), *Erbb2* (58.3%), and *Ret* (46.7%) have the highest presence of novel mutations; while *Braf* and *Egfr* show a small proportion of them (20 and 13.4%, respectively; Supplementary File 1).

The oncogenic driver potential of these novel mutations was assessed using the Cancer Genome Interpreter engine OncodriveMUT ^25^. The analysis revealed that 66.1% of these novel mutations are indeed Tier 1 predicted driver mutations and 15.1% are Tier 2 predicted driver mutations (Supplementary File 2). These are present across nearly all the tested genes (Figure 1C). *Fgfr4, Fgfr3, Met, Mtor and Ros1* are genes with a relevant proportion of novel potentially driver mutations. Regarding the five most frequently mutated genes in the cohort (Figure 3), most novel potentially driver mutations are observed in *Met* and *Alk*. Focusing in NSCLC actionable genes harboring recurrent novel mutations (Table 3 and Supplementary Figure 4) *Alk* displays 13 novel driver mutations, 10 singletons and one recurrent (3 cases of G1537R). For *Met*, 3 mutations are recurrent (T992I, H1094Y and D1010N) and 14 are singletons. Ret displays 5 recurrent mutations and 6 singletons. *Ros1* shows 7 recurrent mutations and other 12 singletons and *Errb2* one recurrent and 17 singletons. Most of the novel variants are located right at/or near the phospho-tyrosine kinase domain (Supplementary Figure 4).

To understand the associations between patient level characteristics and the clinical utility of genomic variants, a radar plot analysis for actionable, likely actionable, known and novel variants subgroups was performed (Figure 3B). A relevant set of novels predicted oncogenic variants is used as a reference (gray shade area in each plot). The analysis shows that gender, age, non-smoker status, biopsy type and predicted pathogenicity traits (CADD) had the same fraction between this subgroup of novel mutations compared to Actionable or Likely-Actionable variants (Figure 3B), while the novel subgroup shows a higher proportion of family history of cancer (FHOC).

## Discussion

The primary goal of this study is to describe the occurrence of known and novel mutations in NSCLC actionable genes in patients from Chile, Peru and Brazil, representing the Latin American continent. The under-representation of these populations in cancer genomic databases, limit the conclusions that can be drawn about NSCLC targeted treatment on the continent. A total of 1732 subjects were included. While their demographic and pathology characteristics are similar to other patient cohorts, but this group is particularly enriched in Hispanics and ethnically admixed subjects, contributing to reduce the genomic disparities observed in current databases^32, 33^ and to support efforts to improve molecular diagnosis and treatment in Latin America.

The prevalence of *Egfr* actionable and known mutations observed in the NIRVANA cohort (15% in Brazil, 12% in Chile, 24% in Peru, Figure 2B) is slightly inferior to observed in CLICaP study by Arrieta et al. ^11^ for Argentina (14.4%), and significantly inferior to the observed in Peru (51.1%). NIRVANA *Egfr* prevalence in Brazil is significantly inferior as observed by Andreis et al. ^13^ (19.2%) and to what was observed in Latinos by Lopez-Chavez et al. (23.3%) and from Peru (37%), but similar to observations made in Venezuela (10%) or Bolivia (7.7%) ^15^. *Egfr* mutations in CLICaP study were assessed using qPCR and limited to exons 19 and 21, so a direct comparison with NGS is not straightforward. NGS was used by Andreis et al. ^13^ but they utilized an AmpliSeq custom panel (Thermo Fisher Scientific) to identify somatic mutations and a different reference sequence (NM_005228.3). Lopez-Chavez et al. used PCR and pyrosequencing limited to exons 19-21. For Alk, in Brazil, Andreis et al. reported a frequency of 4% ^13^, inferior to our study when estimated by Ventana Alk (D5F3). Arrieta et al. ^12^ reported, using *Alk* FISH and focused on t(2;5), a higher prevalence in Brazil (6.8%), and also in Chile (8.6%) and in Peru (10.8%). For *Ros1*, the 28 novel mutations (Table 3) could play an important role in underestimation of its prevalence, increasing it from 1.5% to 3% in Brazil (Supplementary Figure 1B) and from 1.4% to 2% in Chile (Supplementary Figure 1A), but still far from the 6.3% observed in TCGA study by Campbell et al.^34^. These analysis reveals interesting and significant differences in biomarkers prevalence across different Latin America countries, portraying the need to study biomarkers epidemiology at the country level to properly develop public health policies for NSCLC treatments, but also reveals the need of further research to understand the impact of environment and genetics on biomarker prevalence in admixed patients populations.

**Table 3.** Novel variants. Single nucleotide variants counts. Only OncoKB Tier 1 and Tier 2 where considered. Variants that occur less than 2 times were aggregated as *Others*.

| TABLE 3. Novel variants (SNV) |  |
| --- | --- |
| ***Alk*** | n |
| G1537R | 3 |
| Others | 10 |
| ***Met*** | n |
| T992I | 9 |
| H1094Y | 4 |
| D1010N | 2 |
| Others | 14 |
| ***Ret*** |  |
| E902K | 3 |
| A877V | 2 |
| S909N | 2 |
| V882I | 2 |
| V899I | 2 |
| Others | 6 |
| ***Ros1*** |  |
| G1969E | 3 |
| G1954R | 3 |
| G1957E | 2 |
| G1973R | 2 |
| M2029I | 2 |
| P2007L | 2 |
| V1977I | 2 |
| Others | 12 |
| ***ErbB2*** |  |
| A879T | 2 |
| Others | 17 |
| # singleton in other genes | 298 |

Even though it is not part of the present work, the rate of successful nucleic acid extraction and sequencing processes is below to what is commonly reported in clinical studies, reaching only to 60% on RNA and 50% on DNA samples, potentially impacting or adding bias to the prevalence estimations. This may be due to the nature of the samples, as these were collected following real-world conditions and eligibility criteria, isolating nucleic acids from small FFPE tissue pieces^35, 36^ and cellularity levels as low as 5%. These parameters represented the most common fixation method used in South America and the complexity often faced by clinicopathological diagnosis. An alternative to overcome these difficulties is the use of liquid biopsy from plasma ctDNA as an improved diagnostic assay, which had demonstrated a strong genomic alteration match in this cancer type^35^. Further investigation of the relationship between histopathological factors and the quality and amount of nucleic acids extraction could improve the ratio of successful sequencing and hence, contribute to the adoption of this technology to a wider extent.

Regarding factors over clinically actionable or likely-actionable mutations, we observe that *Egfr* occurrences are slightly higher for subjects that have never smoked, and this effect increases if the subjects also had a history of cancer in their family in agreement with the literature ^37^. For *Alk*, female subjects show slightly more *Alk* occurrences than males. This difference is notorious for young women (within 27-35 years old) self-reported as never-smokers, and former-smokers, as previously reported^38-40^. Additionally, we observed that young women self-reported as never-smokers and Native Amerindian display a significantly higher occurrence of Alk mutations.

### Novel mutations

Thanks to a global effort, the knowledge on cancer somatic mutations grows daily. COSMIC, the Catalogue of Somatic Mutations in Cancer^41^, is the world’s largest and most comprehensive resource for exploring the impact of somatic mutations in human cancer. To provide a perspective, the number of coding mutations grew from 9,733,455 in 1,412,466 samples back in September 2019 to 11,453,569 mutations in 1,443,198 samples in April 2020. So, it is very likely that today novel somatic variations will become known somatic mutations in the near future. But the real challenge with novel, rare or infrequent mutations is related to precision medicine and targeted therapies prescription labelling, since clinical trials and the subsequent regulatory approval are tight to the specific mutations that were assayed and considered in the trial. On the other hand, novel, rare or infrequent mutations are not active subjects of research, neither at the clinical trial or cancer genomics level, so evidence about their oncogenic driver role builds up slowly. Many times, treating physicians facing a patient harboring a novel, rare or infrequent mutation, will face uncertainty and the possibility of a lost real therapeutic opportunity and a less than ideal treatment scheme.

The analysis of NIRVANA subjects shows that 1714 mutations were detected. Actionable, likely actionable and known mutations account for 63.5% of these and novel mutations represent 36.5% of the total number of mutations. Following the Cancer Genome Interpreter analysis, 67.3% of the novel mutations are classified as Tier 1 and 0.8% as Tier 2 driver mutations. If we focus the analysis in NSCLC actionable gene ^21^, like *Egfr, Alk, Ros1, Erbb2, Met, Ret* and *Braf*, all combined display 427 actionable mutations in the whole cohort. This number could increase to 512 (a 120% increase) if the 85 novel Tier 1 driver mutations were included. Nonetheless, Cancer Genome Interpreter annotations about deleterious effects, oncogenic classification and prescriptions need further validation at the *in vitro* and *in vivo* level to properly assess its clinical impact.

Novel somatic mutations were strictly filtered in terms of coverage, quality and potential clinical significance with a high proportion of Tier 1 findings; still, other elements might be faultily promoting them, such as the underrepresentation of the Latin American population in the current germline reference genome databases. To reduce the rate of germline-somatic misclassification, alternative variant calling methods are under development for low cellularity and lack of cancer-normal matched DNA samples^42^. Nonetheless, the discovery of novel germline mutations could also open new therapeutics opportunities for these patients. In fact, subjects harboring novel mutations have a higher proportion of family history of cancer and the predicted drivers show a pathogenicity score (CADD) similar to the actual actionable mutations (Figure 3B).

In summary, in this work a large number of Latin American subjects was analyzed to obtain a deep description of mutations present in NSCLC patients. The occurrence of novel somatic mutations could be evidence of genetic differences between reference and Latin-American patients. This more comprehensive and complete panorama of genomic mutations in Brazil, Chile and Peru should open the discussion about current and novel drug recommendations for NSCLC patients in Latin America.

## Data Availability

All data generated or analysed during this study are included in this published article (and its supplementary information files).

## Acknowledgments

The authors thank the patients who consented to provide tumor material and clinical data that was used in this study. Authors thank the Institutional BioBank of the A.C. Camargo Cancer Center, Sao Paulo, Brazil. ED-N is a research fellow from Conselho Nacional de Desenvolvimento Científico e Tecnológico (CNPq), Brazil. The NIRVANA team includes the following Investigators, Institutions, central laboratories and its Researchers:

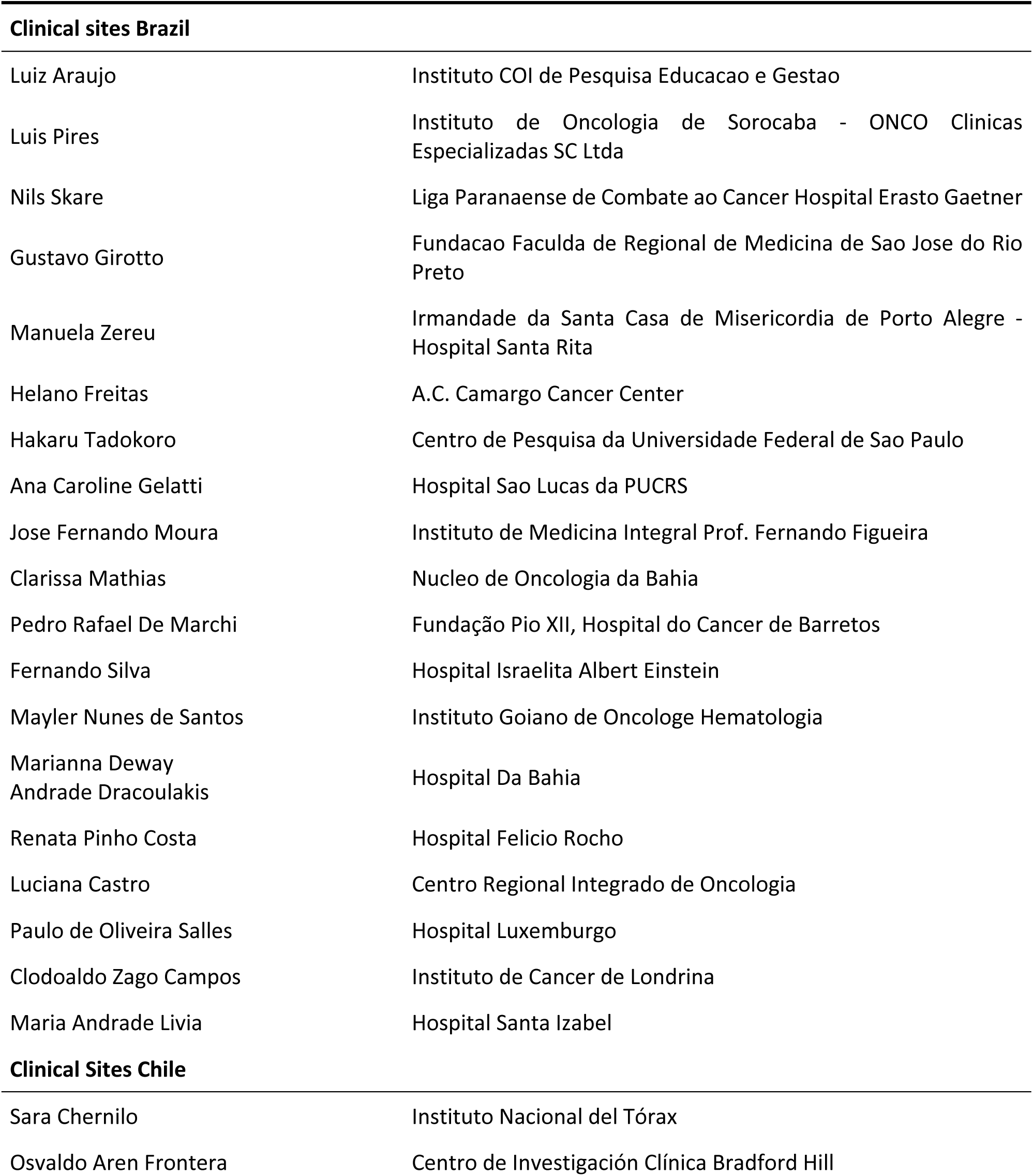
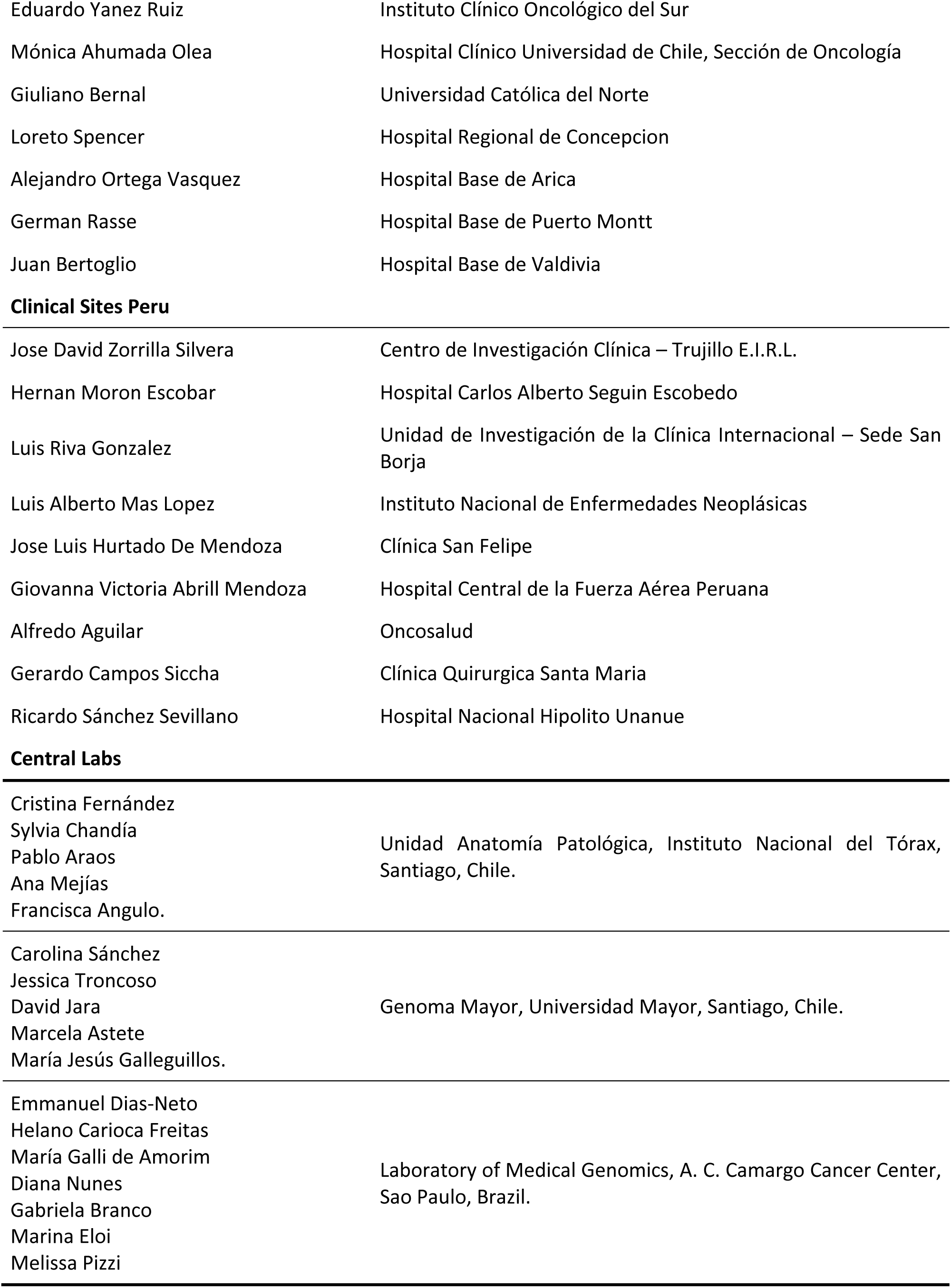
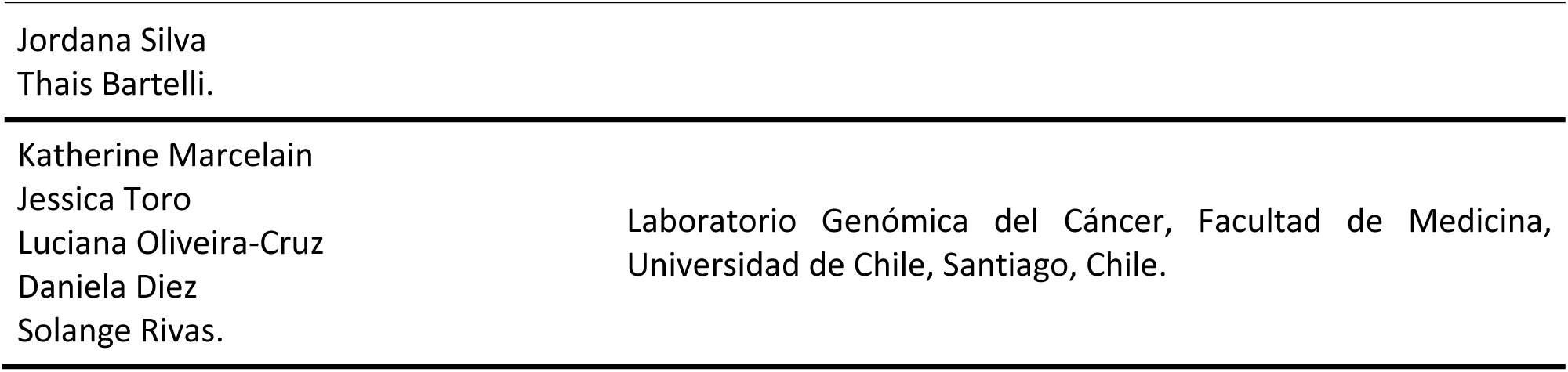

## Conflict of interest statement

GS, AB, RA, RAC, MF, LR, DA, RL, JC and PP were Pfizer Chile employees. HF, EDN, DNN, GPB, MGA, CF, GB, JF, MA, SC, OA, MLS, GR, CS, KM and SR received a grant and non-financial support for to perform this work for CEMP Pfizer Chile. Outside this work, HF discloses personal fees and non-financial support from Pfizer and BMS and non-financial support from AstraZeneca and Roche.

## Funding

This work received support from the Chilean Government CORFO International Center of Excellence Program (Grant # 13CEE2-21602) and Thermo Fisher Scientific. The funding sources were not involved in the study design, the collection, analysis and interpretation of data; in the writing of the report, neither in the decision to submit the article for publication.

## Author Contributions

- **Conceptualization:** Gonzalo Sepúlveda-Hermosilla, Alejandro Blanco, Matías Freire, Paola Pérez, Emmanuel Dias-Neto, Helano Freitas, Rodrigo Assar and Ricardo Armisén.
- **Data acquisition and curation:** Gonzalo Sepúlveda-Hermosilla, Alejandro Blanco, Rodrigo Assar, Matias Freire, Paola Pérez, Carolina Sánchez, Cristina Fernández, Diana Noronha Nunes, Diego Ampuero, Emmanuel Dias-Neto, Germán Rasse, Mónica Ahumada, Giuliano Bernal, Jacqueline Flores, Helano Freitas, Javier Cáceres, Katherine Marcelain, Liliana Ramos, Maria Galli de Amorim, Gabriela Pereira Branco, María Loreto Spencer, Osvaldo Aren, Rodrigo Lizana, Sara Chernilo, Solange Rivas and Ricardo Armisen.
- **Formal analysis:** Gonzalo Sepúlveda-Hermosilla, Rodrigo Assar, Alejandro Blanco, Javier Cáceres and Ricardo Armisen.
- **Methodology:** Rodrigo Assar, Gonzalo Sepúlveda-Hermosilla, Alejandro Blanco, and Ricardo Armisen.
- **Writing - original draft:** Rodrigo Assar, Gonzalo Sepúlveda-Hermosilla, Alejandro Blanco, Matías Freire and Ricardo Armisen.
- **Writing - review & editing:** all co-authors and Ricardo Armisén.

**Abbreviations**
NGS: Next-generation Sequencing
NSCLC: Non-small cell lung cancer
OFA: Oncomine Focus Assay
VEP: Variant Effect Predictor
COSMIC: Catalog of Somatic Mutations in Cancer
VCF: Variant Calling File
TCGA: The Cancer Genome Atlas
ICGC: International Cancer Genome Consortium
SNV: Single Nucleotide Variant
FHOC: Family history of cancer
TKI: tyrosine kinase inhibitors.

## SUPPLEMENTARY FILES LEGENDS

**Supplementary File 1**. Statistics of mutation occurrences per level of evidence and country. Sheet1: Number of occurrences (percentage) of each observed mutation at each country and Fisher Exact test OR and p-value of enrichment. P-values shown only if the total number of occurrences is bigger than 3. Sheet 2: Number of markers obtained by type of mutation (level of evidence), and percentage of markers being Novel.

**Supplementary File 2**. Data listings. Demographic-clinical data and Genomic results. Sheet1: Demographical-clinical data. Sheet2: Annotated results of DNA variants and RNA fusions.

**Supplementary Figure 1.**
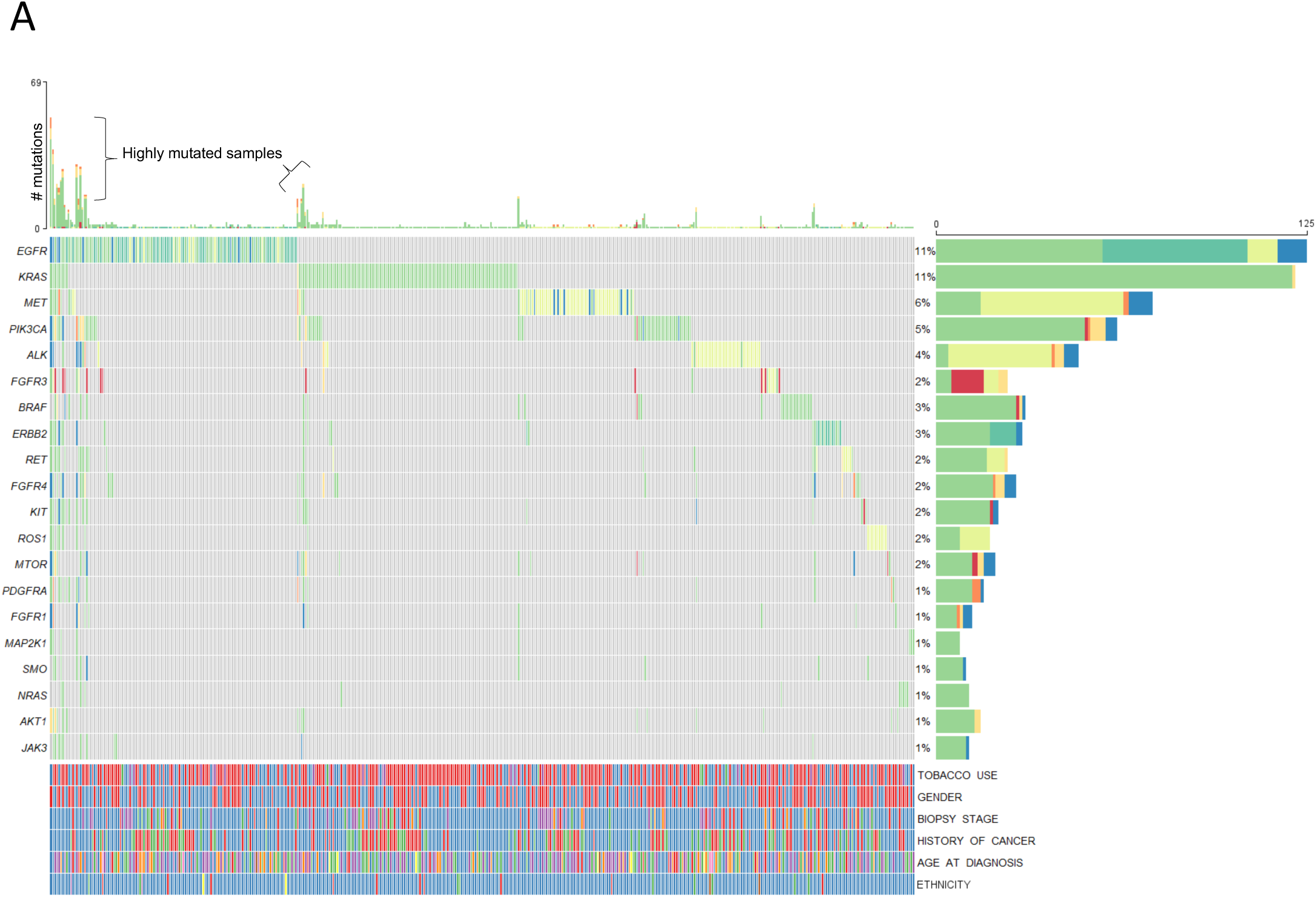
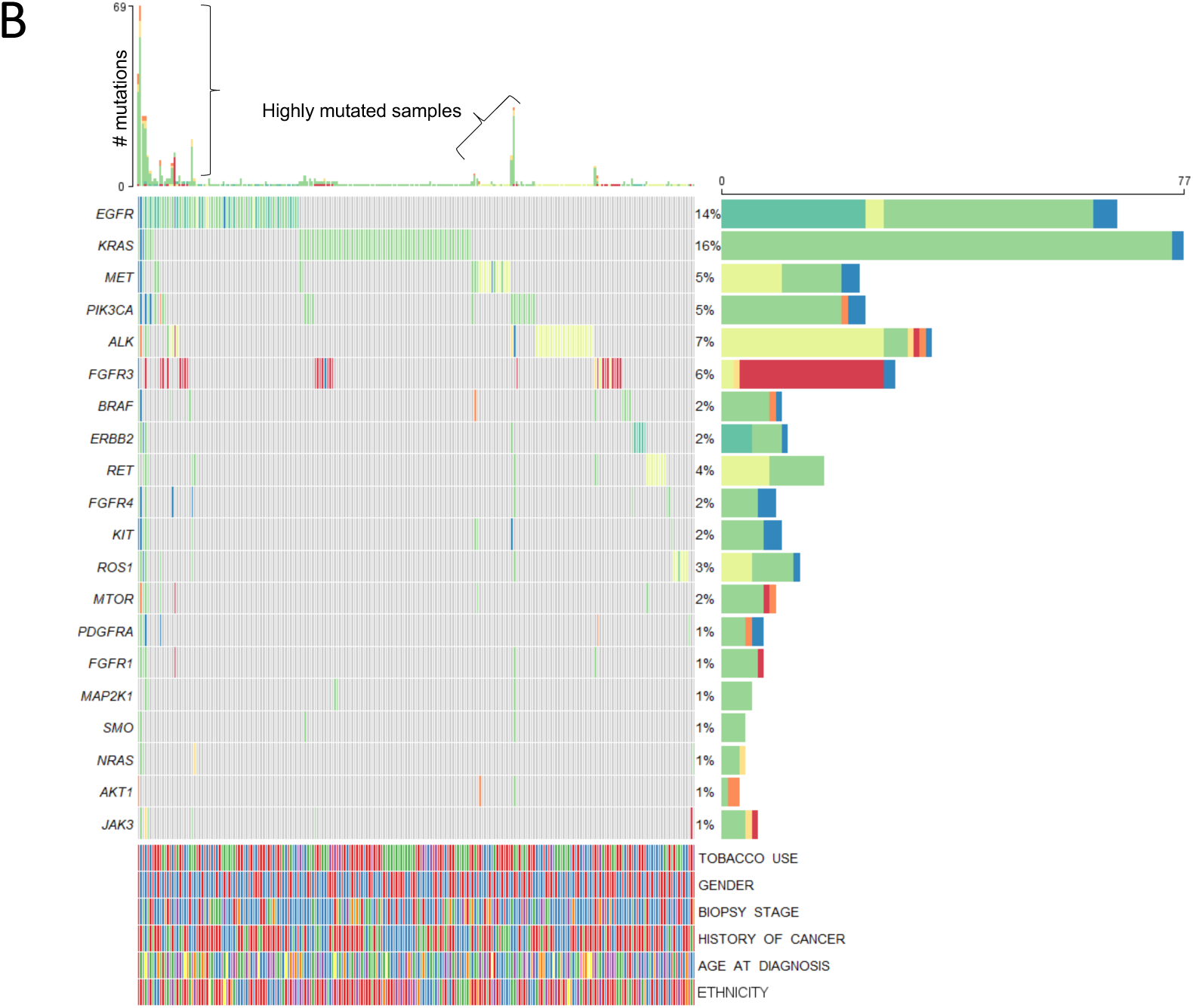
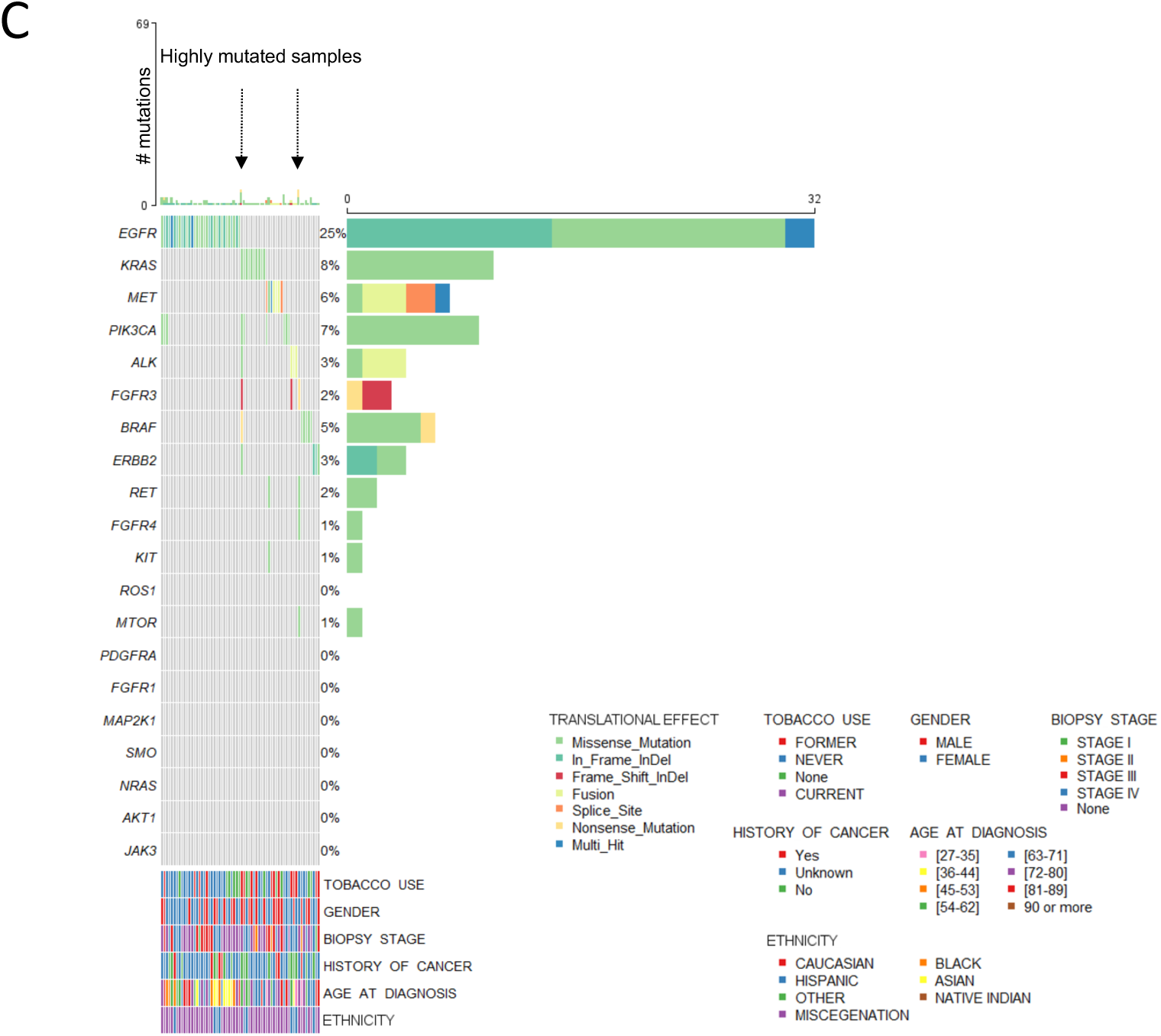
Comprehensive Oncoprint representation of all alterations in the cohort. Rows and columns represent genes and each subject, respectively. Below, additional tracks color-code the most relevant clinical aspects of each subject. Left and upper bars summarize all findings at the gene or subject level, respectively. (A) Oncoprint for Chilean Subjects (B) Oncoprint for Brazilian Subjects. (C) Oncoprint for Peruvian Subjects.

**Supplementary Figure 2.**
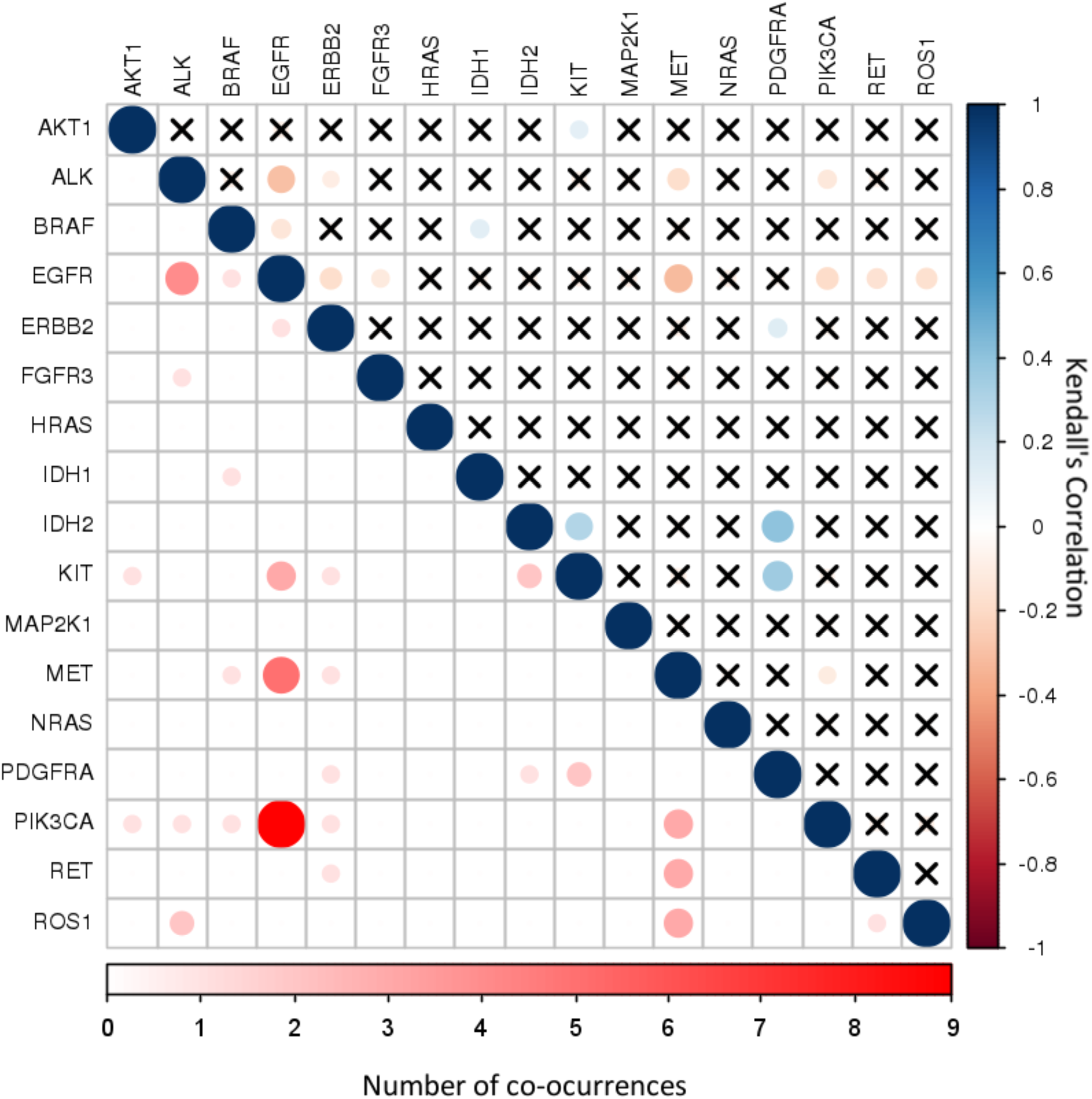
Co-occurrence matrix - occurrences of every pair of NGS-OFA actionable or likely-actionable markers. Right-superior triangle shows Kendall’s correlations and left-inferior triangle the number of co-occurrences. For correlations, only the correlations with a confidence interval of 95% are shown, the non-significant are marked with an “X”. The size and color of the circles represents the strength of the correlation for the upper triangle, and co-occurrences counts for the lower triangle.

**Supplementary Figure 3.**
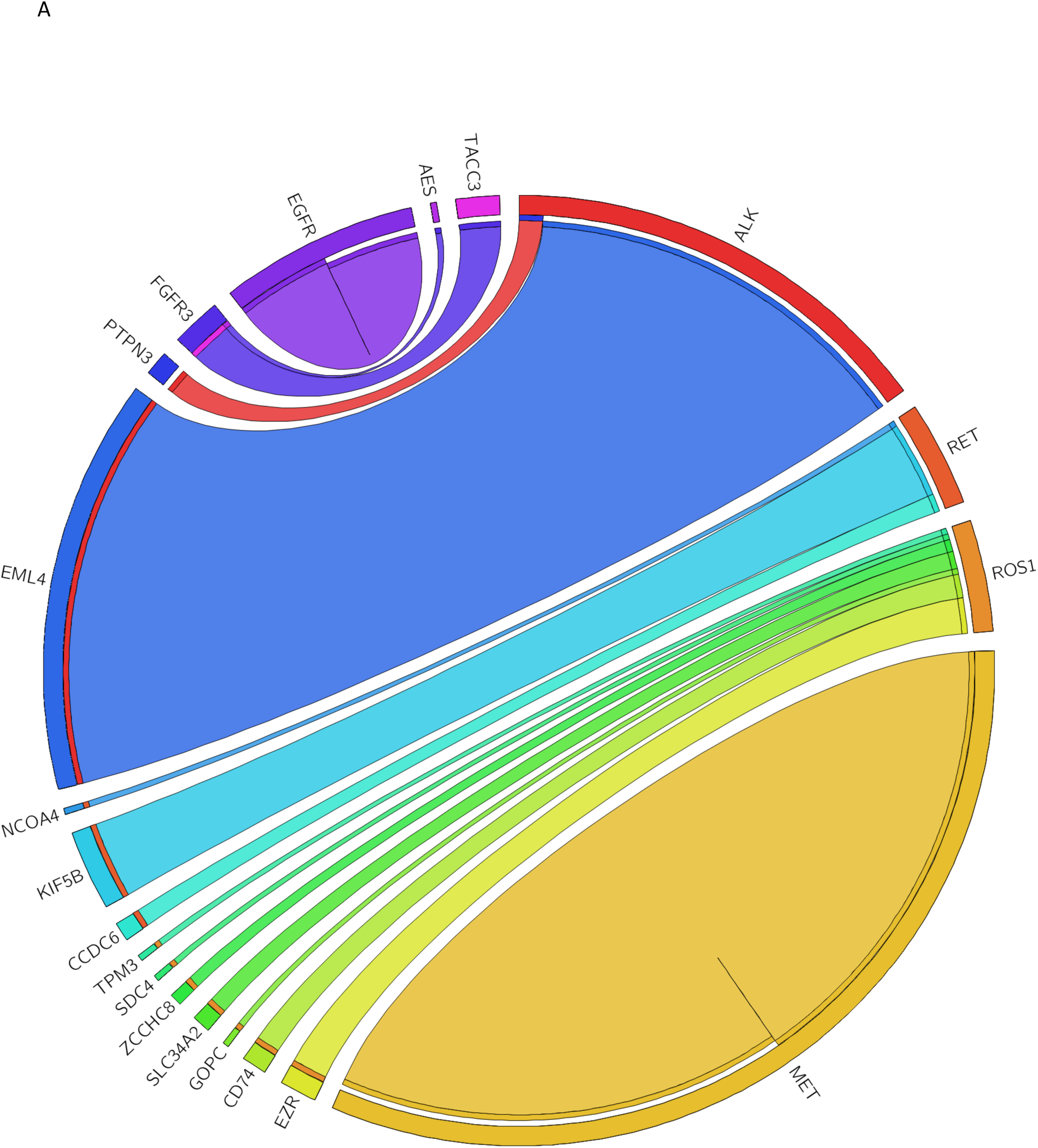
Circos plot. The RNA fusions detected by breakpoint assay are shown as a circus plot. The width of the edges between a pair of genes, represents the proportion observed in the cohort.

**Supplementary Figure 4.**
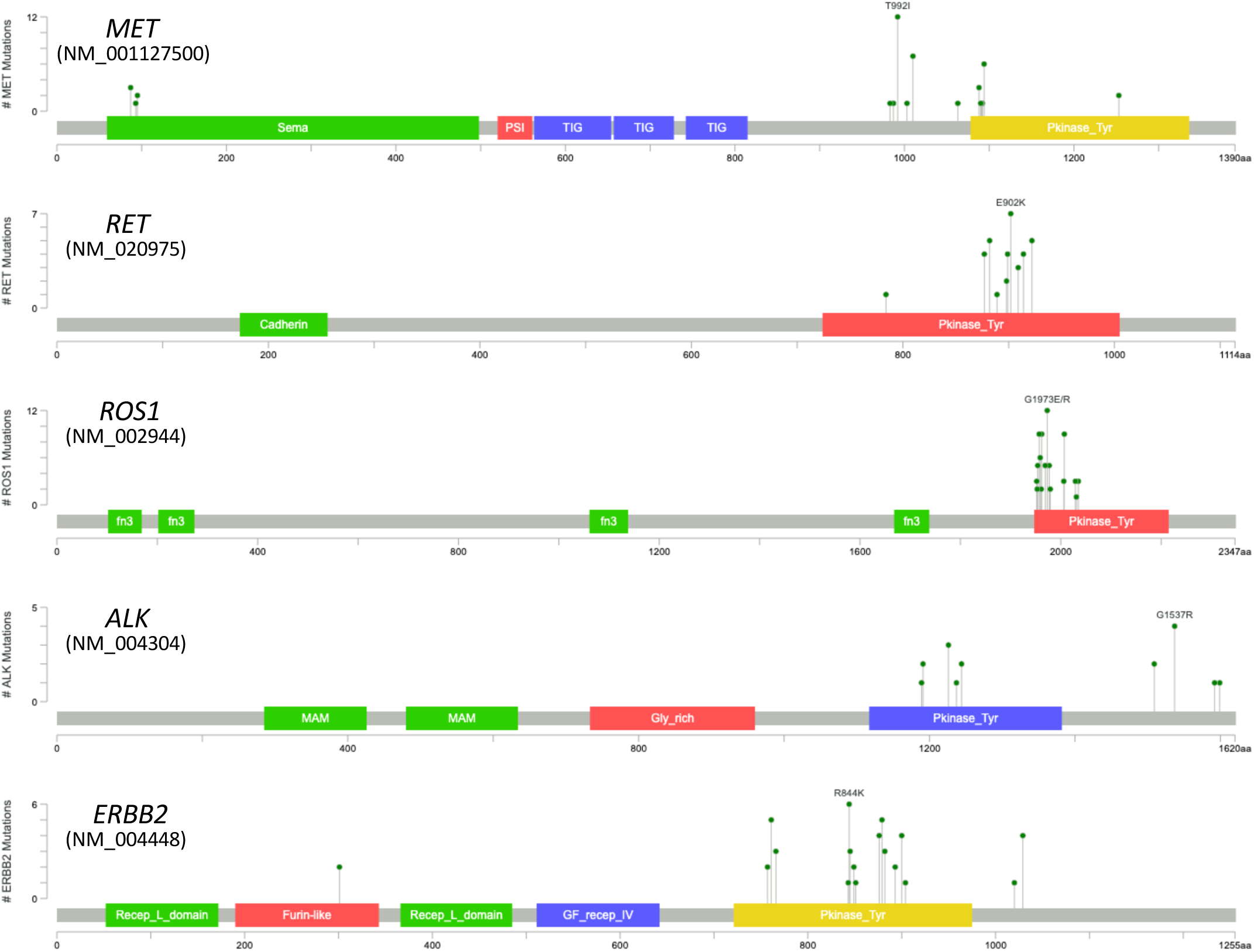
Protein locations and frequency of Novel somatic mutations. Colored boxes represent protein domains at a determined amino acid location (x-axis). Hight of the green colored dots demarks the frequency of each mutation (y-axis). Labeled variants are the most frequent in each gene.

**Supplementary Table 1.** Logistic Regression results analysis. OR value and significance (p-value) over occurrences of *Egfr, Alk*, and *Ros1*. Dashes denote not significant p-values, while the significant ones were ranked as: ** ≤ 0.001; *≤ 0.01

| **Factor** | ***Egfr*** | ***Alk*** | ***Ros1*** |
| --- | --- | --- | --- |
| *Never smoker* | 1.26 ** | - | - |
| *Former smoker* | - | - | - |
| *Hispanic ethnicity* | 1.17 ** | - | 1.03 * |
| *Caucasian ethnicity* | 1.10 ** | - | 1.04 * |
| *Female* | - | 1.03 * | - |
| *Native Indian* | - | 1.54 ** | - |
| *Caucasian ethnicity* | - | 1.06 * | - |
| *Never smoker with family history of cancer* | 1.34 ** | - | - |
| *Never smoker with 27-35 years old* | - | 1.35 * | - |
| *Former smoker with 27-35 years old* | - | 2.72 ** | - |
| *Native Indian never smoker with 27-35 years old* | - | 2.72 ** | - |

## Notes

### Clinical Trial

NCT03220230

### Author Declarations

CONEP SEPN 510 Norte- Bloco A- 3DEGREE Andar- Edificio Ex-INAN, Unidade II - Ministerio da Saude Brasilia, DF 70750-521 BRAZIL Comite Etico Cientifico Av. Salvador 364, Servicio de Salud Metropolitano Oriente Providencia, SANTIAGO, RM 7500922 CHILE Comite de Etica Hospital Clinico Universidad de Chile Avenida Santos Dumont 999 Santiago, RM 8380456 CHILE Comite Etico Cientifico. Servicio de Salud Concepcion San Martin 1436 Concepcion, REGION DEL BIO BIO 4070038 CHILE Comite Institucional de Bioetica de VIA LIBRE(CIB) Jr. Paraguay 490, Cercado de Lima Lima, LIMA, PERU 01 PERU Comite de Etica en Investigacion del INEN Av. Angamos Este 2520, Surquillo 15038 Lima PERU Comite Institucional de Bioetica Via Libre Jr Paraguay 490 Lima PERU Comite Institucional de Etica en Investigacion Hospital Nacional Hipolito UNANUE Av. Cesar Vallejo 1390, El Agustino Lima PERU

## References

1. WHO. Latest global cancer data: Cancer burden rises to 18.1 million new cases and 9.6 million cancer deaths in 2018. 2018.

2. Stewart BWW, C. P. World Health Organization: Geneva. World Cancer Report 2014.

3. Rafiemanesh H, Mehtarpour M, Khani F, et al. Epidemiology, incidence and mortality of lung cancer and their relationship with the development index in the world. J Thorac Dis 2016;8:1094–1102.

4. Sandler A, Gray R, Perry MC, et al. Paclitaxel-carboplatin alone or with bevacizumab for non-small-cell lung cancer. N Engl J Med 2006;355:2542–2550.

5. Shepherd FA, Rodrigues Pereira J, Ciuleanu T, et al. Erlotinib in previously treated nonsmall-cell lung cancer. New England journal of medicine 2005;353:123–132.

6. Schmidt KT, Chau CH, Price DK, et al. Precision Oncology Medicine: The Clinical Relevance of Patient Specific Biomarkers Used to Optimize Cancer Treatment. J Clin Pharmacol 2017;56:1484–1499.

7. Antonicelli A, Cafarotti S, Indini A, et al. EGFR-targeted therapy for non-small cell lung cancer: focus on EGFR oncogenic mutation. Int J Med Sci 2013;10:320–330.

8. Doherty M, Metcalfe T, Guardino E, et al. Precision medicine and oncology: an overview of the opportunities presented by next-generation sequencing and big data and the challenges posed to conventional drug development and regulatory approval pathways. Ann Oncol 2016;27:1644–1646.

9. Gainor JF, Varghese AM, Ou SI, et al. ALK rearrangements are mutually exclusive with mutations in EGFR or KRAS: An analysis of 1,683 patients with non-small cell lung cancer. Clin Cancer Res 2013;19.

10. Bergethon K, Shaw AT, Ou SH, et al. ROS1 rearrangements define a unique molecular class of lung cancers. J Clin Oncol 2012;30:863–870.

11. Arrieta O, Cardona AF, Martin C, et al. Updated Frequency of EGFR and KRAS Mutations in NonSmall-Cell Lung Cancer in Latin America: The Latin-American Consortium for the Investigation of Lung Cancer (CLICaP). J Thorac Oncol 2015;10:838–843.

12. Arrieta O, Cardona AF, Bramuglia G, et al. Molecular Epidemiology of ALK Rearrangements in Advanced Lung Adenocarcinoma in Latin America. Oncology 2019;96:207–216.

13. Andreis TF, Correa BS, Vianna FS, et al. Analysis of Predictive Biomarkers in Patients With Lung Adenocarcinoma From Southern Brazil Reveals a Distinct Profile From Other Regions of the Country Journal of Global Oncology 2019:1-9.

14. Lopes LF, Bacchi CE. Anaplastic lymphoma kinase gene rearrangement in non-small-cell lung cancer in a Brazilian population. Clinics (Sao Paulo) 2012;67:845–847.

15. Lopez-Chavez A, Thomas A, Evbuomwan MO, et al. EGFR Mutations in Latinos From the United States and Latin America Journal of Global Oncology 2016;2:259–267.

16. Spratt DE, Chan T, Waldron L. Racial/Ethnic disparities in genomic sequencing. JAMA Oncology 2016;2:1070–1074.

17. Yuan J, Hu Z, Mahal BA, et al. Integrated Analysis of Genetic Ancestry and Genomic Alterations across cancers. Cancer Cell 2018;34:549–560.e549.

18. DESA. World Population Prospects 2019..

19. Kettering MS. Available at https://github.com/mskcc/vcf2maf.

20. McLaren W, Gil L, Hunt SE, et al. The Ensembl Variant Effect Predictor. Genome Biol 2016;17:122.

21. Chakravarty D, Gao J, Phillips SM, et al. OncoKB: A Precision Oncology Knowledge Base. JCO Precis Oncol 2017;2017.

22. Tate JG, Bamford S, Jubb HC, et al. COSMIC: the Catalogue Of Somatic Mutations In Cancer. Nucleic Acids Res 2019;47:D941-D947.

23. Cerami E, Gao J, Dogrusoz U, et al. The cBio cancer genomics portal: an open platform for exploring multidimensional cancer genomics data. Cancer Discov 2012;2:401–404.

24. IRB. Cancer Genome Interpreter. Available at https://www.cancergenomeinterpreter.org/home.

25. Tamborero D, Rubio-Perez C, Deu-Pons J, et al. Cancer Genome Interpreter annotates the biological and clinical relevance of tumor alterations. Genome Med 2018;10:25.

26. Li MM, Datto M, Duncavage EJ, et al. Standards and Guidelines for the Interpretation and Reporting of Sequence Variants in Cancer: A Joint Consensus Recommendation of the Association for Molecular Pathology, American Society of Clinical Oncology, and College of American Pathologists. J Mol Diagn 2017;19:4–23.

27. Shirts BH, Konnick EQ, Upham S, et al. Using Somatic Mutations from Tumors to Classify Variants in Mismatch Repair Genes. Am J Hum Genet 2018;103:19–29.

28. Mayakonda A, Lin DC, Assenov Y, et al. Maftools: efficient and comprehensive analysis of somatic variants in cancer. Genome Research 2018;28:1747–1756.

29. Krzywinski. Tableviewer. Available at http://mkweb.bcgsc.ca/tableviewer/.

30. Agresti A, Coull BA. Approximate is better than ‘exact’ for interval estimation of binomial proportions. The American Statistician 1998;52:119–126.

31. Jordan EJ, Kim HR, Arcila ME, et al. Prospective Comprehensive Molecular Characterization of Lung Adenocarcinomas for Efficient Patient Matching to Approved and Emerging Therapies. Cancer Discov 2017;7:596–609.

32. Landry LG, Ali N, Williams DR, et al. Lack Of Diversity In Genomic Databases Is A Barrier To Translating Precision Medicine Research Into Practice. Health Aff (Millwood) 2018;37:780–785.

33. Sirugo G, Williams SM, Tishkoff SA. The Missing Diversity in Human Genetic Studies. Cell 2019;177:26–31.

34. Campbell JD, Alexandrov A, Kim J, et al. Distinct patterns of somatic genome alterations in lung adenocarcinomas and squamous cell carcinomas. Nature Genetics 2016;48:607–616.

35. Yang H, Zhang J, Zhang L, et al. Comprehensive analysis of genomic alterations detected by next-generation sequencing-based tissue and circulating tumor DNA assays in Chinese patients with non-small cell lung cancer. Oncology Letters 2019;18:4762–4770.

36. Kris MG, Johnson BE, Berry LD, et al. Using multiplexed assays of oncogenic drivers in lung cancers to select targeted drugs. JAMA 2014;311:1998–2006.

37. Sun J-M, Lira M, Pandya K, et al. Clinical characteristics associated with ALK rearrangements in never-smokers with pulmonary adenocarcinoma. Lung Cancer 2014;83:259–264.

38. Shaw AT, Yeap BY, Mino-Kenudson M, et al. Clinical features and outcome of patients with non-small-cell lung cancer who harbor EML4-ALK. J Clin Oncol 2009;27:4247–4253.

39. Lee YJ, Kim JH, Kim SK, et al. Lung cancer in never smokers: change of a mindset in the molecular era. Lung Cancer 2011;72:9–15.

40. Okazaki I, Ishikawa S, Ando W, et al. Lung Adenocarcinoma in Never Smokers: Problems of Primary Prevention from Aspects of Susceptible Genes and Carcinogens. Anticancer Research 2016;36:6207–6224.

41. Bamford S, Dawson E, Forbes S, et al. The COSMIC (Catalogue of Somatic Mutations in Cancer) database and website. Br J Cancer 2004;91:355–358.

42. Sukhai MA, Misyura M, Thomas M, et al. Somatic Tumor Variant Filtration Strategies to Optimize Tumor-Only Molecular Profiling Using Targeted Next-Generation Sequencing Panels. J Mol Diagn 2019;21:261–273.

